# Effect of heterologous vaccination regimen with Ad5-nCoV CanSinoBio and BNT162b2 Pfizer in SARS-CoV-2 IgG antibodies titers

**DOI:** 10.1101/2021.10.07.21264657

**Authors:** Maria Elena Romero-Ibarguengoitia, Arnulfo González Cantú, Yodira Guadalupe Hernández-Ruíz, Ana Gabriela Amerndariz-Vázquez, Diego Rivera-Salinas, Laura Patricia Montelongo-Cruz, Gerardo Francisco del Río-Parra, Irene Antonieta Barco-Flores, Rosalinda González-Facio, Miguel Ángel Sanz-Sánchez

## Abstract

**Introduction:** The efficacy with one dose Ad5-nCoV has been concerned. As a result, some patients have self-reported getting a boost with BNT. Therefore, this study aimed to compare SARS-CoV-2 spike 1-2 IgG antibodies in plasma samples between two groups: one group immunized with Ad5-nCoV and another with a heterologous vaccination regimen with Ad5-nCoV and BNT.

**Methods:** Prospective observational study included a subgroup analysis of patients who received the Ad5-nCoV immunization during the first trimester of 2021 in a Northern city of Mexico; and agreed to a follow-up for an entire year through SARS-CoV-2 specific IgG antibodies measurement samples. During the three months follow-up, some patients self-reported receiving a BNT boost. We report IgG levels from basal, 21-28 days after Ad5-nCoV dose, three months, and an additional 21-28 days after BNT boost.

**Results:** Seventeen patients 40 (16) years old, 52.9% men, were analyzed. We created four groups: (G1) patients vaccinated with Ad5-nCoV with no history of SARS-COV-2 (n=4), (G2) patients vaccinated with Ad5-nCoV and the first shot of BNT with no history of SARS-COV-2 (n=6), (G3) patients vaccinated with Ad5-nCoV with history of SARS-COV-2 (n=5), and (G4) patients vaccinated with Ad5-nCoV and the first shot of BNT with history of SARS-COV-2 (n=2).

The group immunized with a heterologous vaccine scheme reported higher antibodies after 21-28 days of follow-up after BNT boost. Median (IQR): G1 46.7 (-), G2 1077.5 (1901), G3 1158.5 (2673.5), and G4 2090 (-) (p<0.05). Headache was the most frequent adverse reaction when patients received Ad5-nCoV (n = 10, 83%), and pain at the injection site was the most frequent adverse reaction with BNT boost (n = 5, 83.3%).

**Conclusion:** Patients receiving a BNT boost after Ad5-nCoV had higher SARS-CoV-2 spike 1-2 IgG antibodies titers with no severe adverse reaction.

**Author Approval:** all authors read an approved the final version of the manuscript

**Competing Interests:** The authors have declared no competing interest

**Funding:** The research was supported by private funding provided by the hospital. No external funding was used.

**Ethics statement:** Ethics committee/local Institutional Review Board from the school of Medicine from Universidad de Monterrey gave ethical approval: Ref.:26022021-CN-1e-CI

## Introduction

On March 11, 2020, The World Health Organization (WHO) declared the SARS-COV-2 pandemic due to the increasing cases of atypical pneumonia caused by a novel virus agent called Severe Acute Respiratory Syndrome Coronavirus-2 (SARS-CoV-2)[1]. This disease has had an outstandingly overwhelming impact on the health department. Most hospitals and health centres were overloaded due to no definitive and complete treatment against the agent and its highly contagious nature. According to the WHO, there have been 223,022,538 cases and 4,602,882 deaths worldwide, while in Mexico, 3,465,171 cases and 265,420 deaths to date [2].

This worldwide contingency led to various vaccines against SARS-CoV-2 to decrease severe cases and the need for hospitalization. Multiple vaccines are being deployed globally. Nowadays, there are 22 approved vaccines with different mechanisms of action to develop spike-specific IgG antibodies with neutralizing capacity against SARS-CoV-2 [3]. The different designs of SARS-CoV-2 vaccines are: Messenger RNA vaccines (BNT162b2 Pfizer/BioNTech, mRNA-1273 Moderna), adenoviral-vectored vaccines (AZD1222 Oxford/AstraZeneca, Gam-COVID-VAC/Sputnik V, Janssen, Ad5-nCoV CanSinoBio), protein subunit vaccines (NVX-CoV2373 Novavax, Medicago CoVLP), whole-cell inactivated virus vaccines (CoronaVac Sinovac, BBIBP-CorV Sinopharm), and DNA vaccines (INO-4800 and ZyCoV-D) [4,5].

In Mexico, there have been approved nine vaccines against SARS-CoV-2: 1) BNT162b2 Pfizer/BioNTech, 2) AZD1222 Oxford/AstraZeneca, 3) Ad5-nCoV CanSinoBio, 4) CoronaVac Sinovac, 5) mRNA-1273 Moderna, 6) Janssen/Johnson & Johnson Ad26.COV2.5, 7) Gam-COVID-VAC/Sputnik V; 8) Bharat Biotech Covarix, and 9) BBIBP-CorV Sinopharm (Vero Cells) [6]. These vaccines were distributed according to the national vaccination campaign.

The CanSinoBio vaccine, hereafter referred to as Ad5-nCOV, is an adenovirus type 5 vectored SARS-COV-2 vaccine. It was developed in China as a single-shot vaccine, and it has been approved in 9 countries [7]. The phase III trials showed 68.83% and 95.47% efficacy against all symptomatic infections and severe disease after 14 days of vaccination, respectively [8]. The Pfizer/BioNTech, hereafter referred to as BNT, is an mRNA vaccine that requires two shots, 21 days apart from each dose. The vaccine provides 95% of effectiveness against the infection of SARS-COV-2 in patients with the two shots [9]. It has been approved in 98 countries, and the WHO recommends its application worldwide [10].

To date, the new subject of interest is heterologous vaccine regimens against SARS-COV-2. In Germany, an amplified immune response was observed in patients that used the Oxford/AstraZeneca vaccine as prime dose and BNT as a boost than subjects who received the homologous regimen. However, despite the positive results, heterologous vaccination cannot be recommended because of insufficient proof of safety [11].

In Mexico, the population that received the Ad5-nCoV vaccine was concerned about its efficacy. As a result, they started getting a boost with BNT, despite any medical advice. In the second half of 2021, Hospital Clinica Nova (HCN), in San Nicolas de los Garza, Nuevo Leon, Mexico, started investigating SARS-CoV-2 vaccines. Therefore, this study aims to compare SARS-CoV-2 spike 1-2 IgG antibodies in a vaccinated group with one dose of Ad5-nCoV and a second group that self-reported heterologous vaccination regimen with Ad5-nCoV and BNT.

## Methods

It was a prospective observational study that followed the STROBE guidelines at HCN [12]. This study included an analysis of a subgroup of patients who received the Ad5-nCoV immunization during the first trimester of 2021 in Monterrey, Nuevo Leon, Mexico. The study was approved by the local Institutional Review Board (Ref.:26022021-CN-1e-CI) and conducted per The Code of Ethics of the World Medical Association (Declaration of Helsinki) for human experiments. Due to the prospective nature of the study, every patient had to sign an informed consent form to participate.

The patients were recruited according to the established inclusion and exclusion criteria. The inclusion criteria were: individuals of both genders, between the ages of 18 and 100 years, who had signed up the informed consent form and planned to conclude the immunization regimen of any vaccine given by the Mexican National Health System in compliance with the current immunization regimens. The criteria to exclude patients was: if the time range of interest for this study had passed or if they had previously received a SARS-COV-2 vaccine.

The research team contacted the patients for an informative session of the protocol. After explaining the study to each patient, emphasizing the follow-up along an entire year through SARS-CoV-2 specific IgG antibodies measurement samples, all participants provided written informed consent. After reading it and agreeing to be part of the study, the phlebotomists took a plasma sample. The first sample taken was a baseline level before receiving the first dose of any SARS-COV-2 vaccine.

The Mexican National Health System established that the vaccination campaign against SARS-COV-2 would be divided according to age groups or occupation. Therefore, after 21-28 days of receiving prime-boost vaccination, the research team contacted the participants to take the second plasma sample for IgG antibodies measurement. Then, they waited for the date of the second dose of the vaccine, and then in a period of 21-28 days, they were scheduled to take the third IgG antibodies sample. The fourth, fifth and sixth IgG antibody samples were programmed to be taken three, six, and twelve months, respectively, after receiving the second vaccine dose.

Every time the participants attended the antibodies sample, they had to answer a questionnaire. The basal-sample questionnaire aimed to obtain each patient’s medical history and previous SARS-COV-2 infections. The questionnaires applied after the first and second dose of the vaccine aimed to recognize each application’s adverse events and identify a SARS-COV-2 infection after receiving any vaccine dose. After completing the vaccine regimen, the follow-up questionnaires on the fourth, fifth and sixth IgG antibody samples asked about any suspicious or confirmed SARS-COV-2 infections.

For sampling, 10 to 15 ml of blood were required per venipuncture. The taken sample was in sample tubes with ethylenediaminetetraacetic acid (EDTA) as an anticoagulant, stored at −80 ºC. The laboratory personnel used the LIAISON SARS-CoV-2 S1 / S2 IgG antibody detection kit (Italy) to analyze the samples. It used chemiluminescence immunoassay (CLIA) technology to determine the amount of specific anti-S1 and anti-S2 IgG antibodies against SARS-CoV-2 in plasma samples. It had a sensitivity of 97.4% (95% CI, 86.8-99.5) and a specificity of 98.5% (95% CI, 97.5-99.2). The results were reported as follows: <12.0 AU / ml was considered negative, 12.0 to 15.0 AU / ml was indeterminate, and > 15 AU / ml was positive.

In the three-month follow-up, patients who received Ad5-nCoV were concerned about the effectiveness of a single-dose vaccine, so by their own means, they received a BNT boost, and they reported it to the research team. Additionally, another plasma sample was taken from the entire Ad5-nCoV recruited group twenty-one days after applying BNT in all the participants of the subgroup.

We analyzed the following variables: age, gender, the time between the first and second vaccine, medical history including the confirmed SARS-COV-2 diagnosis (confirmed with a nasal swab and serologic tests before to 21-28 days after BNT boost), adverse effects caused by the Ad5-nCoV, and the first shot of BNT. The analyzed biochemical variables were SARS-CoV-2 quantitative antibodies from the basal sample (S1), 21-28 days post-Ad5-nCoV (S2), three-month follow-up after immunization (S3), and 21-28 days post-first BNT-boost (S4).

The researchers reviewed the quality control and the anonymization of the database. Normality assumption was evaluated with the Shapiro Wilk test and frequency histograms. Descriptive statistics such as median, the interquartile range for quantitative variables, frequencies, and percentages for categorical variables, were computed. Kruskal Wallis test was used for group comparison. The statistical program used was SPSS, version 2. The analysis was two-tailed. A p-value < 0.05 was considered statistically significant.

## Results

The recruited participants immunized with Ad5-nCoV were 17, of which 8 received an additional BNT shot.

We divided the data into four groups: (G1) patients vaccinated with Ad5-nCoV with no history of SARS-COV-2 (n=4), (G2) patients vaccinated with Ad5-nCoV and the first shot of BNT with no history of SARS-COV-2 (n=6), (G3) patients vaccinated with Ad5-nCoV with history of SARS-COV-2 (n=5), and (G4) patients vaccinated with Ad5-nCoV and the first shot of BNT with history of SARS-COV-2 (n=2).

The mean (SD) age was 40 (16) years, and most of the participants were men (n=9, 52.9%). Five patients had confirmed SARS-COV-2 infection by PCR test before immunization, and two patients after the first dose of AD5-nCoV and before the BNT boost. The time between the first vaccine and the first shot of the second one was 93 (5) days. The most-reported comorbidity was obesity (n= 7, 17%), followed by hypertension (n=3, 17%). Table 1 shows the medical history reported by the participants on the basal sample questionnaire.

**Table 1.**
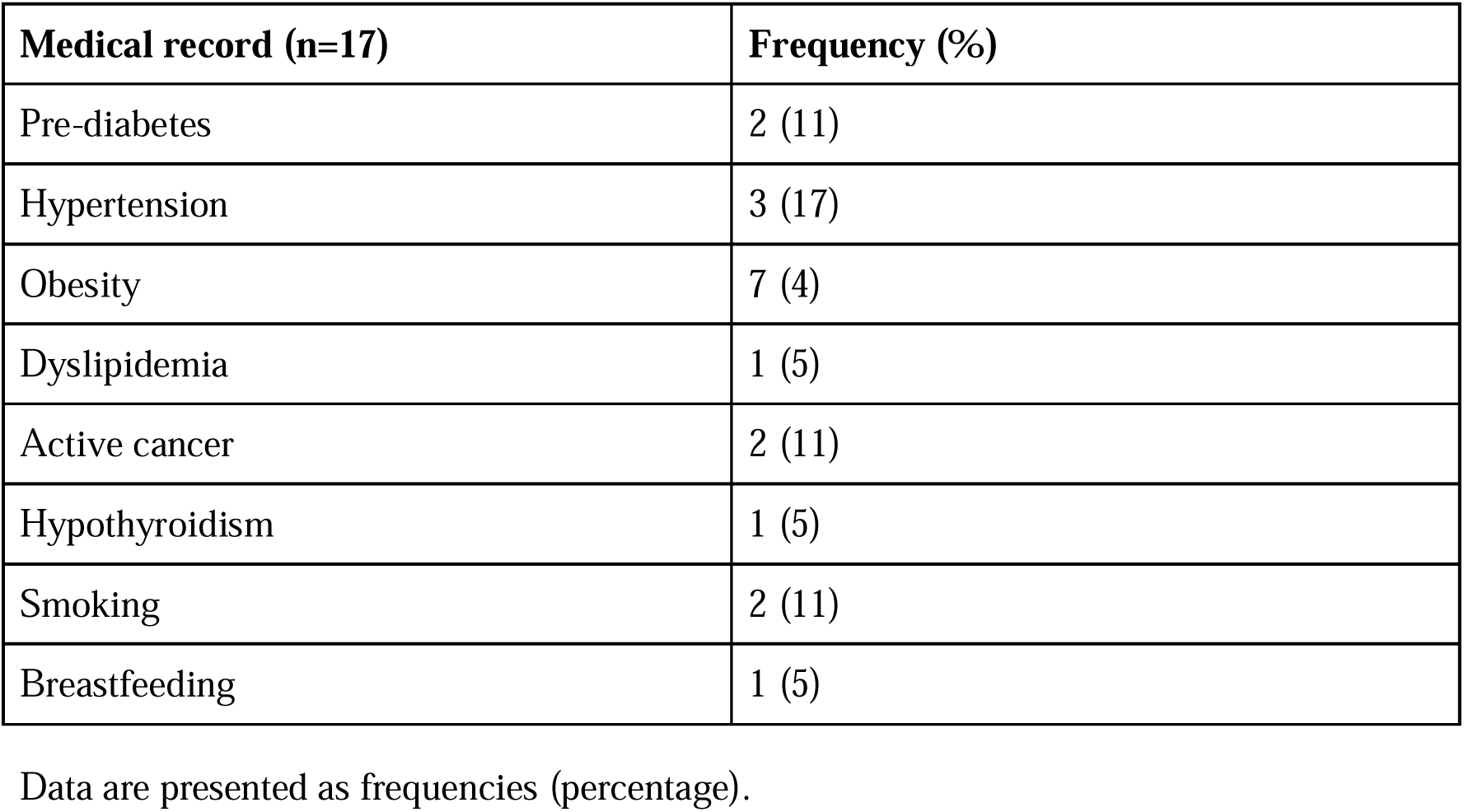
Medical history.

The median baseline (IQR) of the quantitative SARS-CoV-2 spike 1-2 IgG antibody titers obtained from S1 in the four groups were: 3.8 (0), 3.8 (0), 93 (231), and 3.8 (-) (p = 0.030), respectively. The results per group from the S4 were: G1 46.7 (-), G2 1077.5 (1901), G3 1158.5 (2673.5), and G4 2090 (-) (p = 0.050). Table 3 shows the quantitative SARS-CoV-2 spike 1-2 IgG antibody titers against SARS-COV-2 in the different groups of participants depending on the applied vaccines and SARS-COV-2 history.

The most common adverse events related to AD5-nCoV were headache (n=10, 83%), pain at the injection site (n=8,66%), tiredness (n=7, 58%), and their severity was classified as very low (n=7, 58%). The most common adverse events reported after BNT shot were puncture site pain (n=5, 83.3%) and tiredness (n=3, 50%), and their severity was classified as low (n=3, 50%). Table 2 shows the adverse events to vaccinations related to the Ad5-nCoV first shot and the BNT boost shot reported by the participants on the S3 and S4.

**Table 2.**
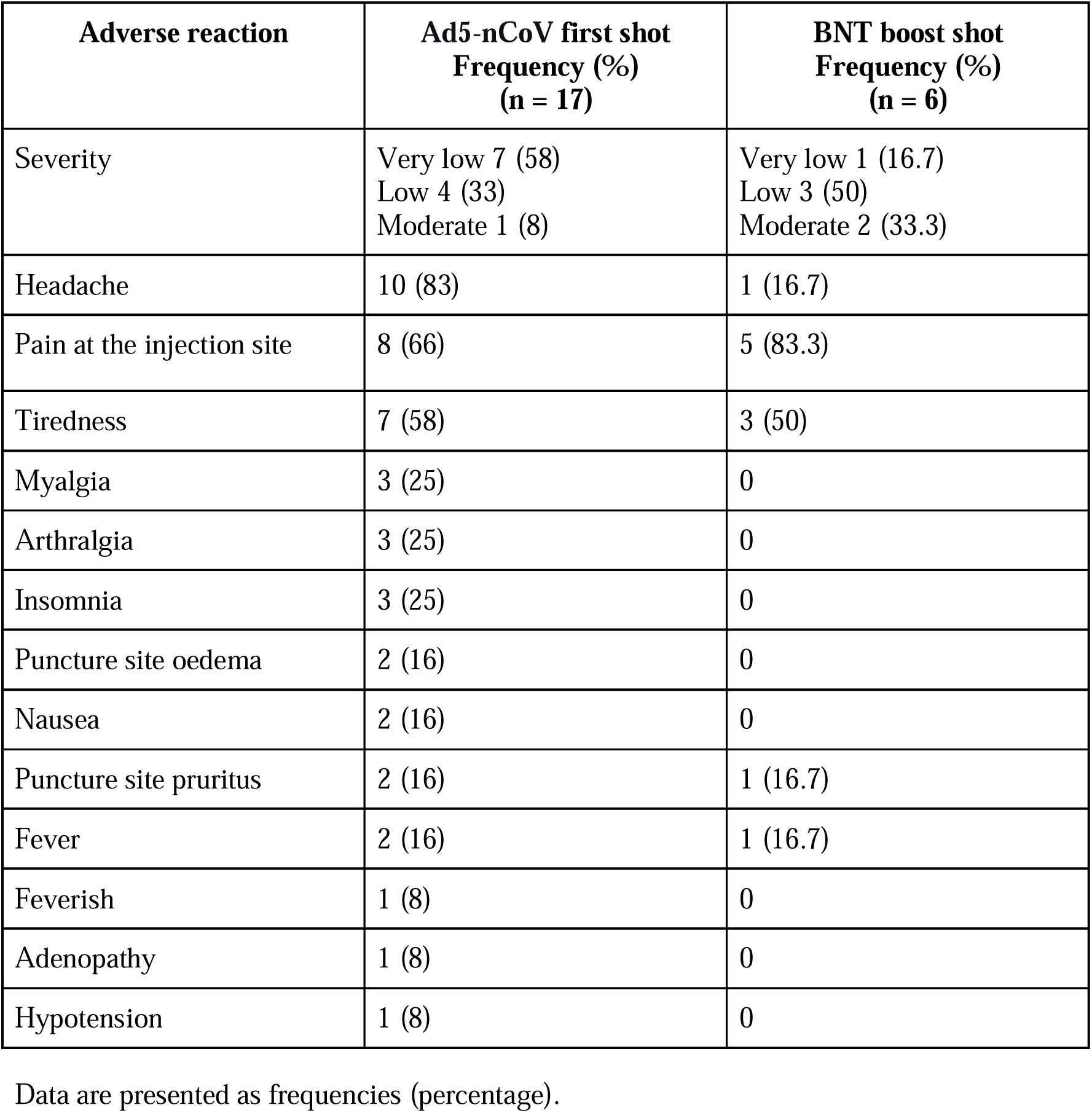
Adverse events in relation to vaccinations.

| **Adverse reaction** | **Ad5-nCoV first shot**<br>**Frequency (%)**<br>**(n = 17)** | **BNT boost shot**<br>**Frequency (%)**<br>**(n = 6)** |
| --- | --- | --- |
| Severity | Very low 7 (58)<br>Low 4 (33)<br>Moderate 1 (8) | Very low 1 (16.7)<br>Low 3 (50)<br>Moderate 2 (33.3) |
| Headache | 10 (83) | 1 (16.7) |
| Pain at the injection site | 8 (66) | 5 (83.3) |
| Tiredness | 7 (58) | 3 (50) |
| Myalgia | 3 (25) | 0 |
| Arthralgia | 3 (25) | 0 |
| Insomnia | 3 (25) | 0 |
| Puncture site oedema | 2 (16) | 0 |
| Nausea | 2 (16) | 0 |
| Puncture site pruritus | 2 (16) | 1 (16.7) |
| Fever | 2 (16) | 1 (16.7) |
| Feverish | 1 (8) | 0 |
| Adenopathy | 1 (8) | 0 |
| Hypotension | 1 (8) | 0 |

Data are presented as frequencies (percentage).

**Table 3.** Quantitative SARS-CoV-2 spike 1-2 IgG antibody titers against SARS-COV-2 in the different groups of participants depending on the applied vaccines and SARS-COV-2 history.

| **Group (n=17)** | **Before the first shot of vaccine (S1) (QIR)** | **21-28 days post Ad5-nCoV (S2) (QIR)** | **3 months after Ad5-nCoV (S3) (QIR)** | **21-28 days post-BNT (S4) (QIR)** |
| --- | --- | --- | --- | --- |
| **Ad5-nCoV/SARS-COV-2 negative (G1) (n = 4)** | 3.8 (0) | 49 (75) | 54 (119) | 46.7 (-) |
| **Ad5-nCoV/BNT/SARS-COV-2 negative (G2) (n = 6)** | 3.8 (0) | 62.25 (77) | 28.6 (73) | 1077.5 (1901) |
| **Ad5-nCoV/SARS-COV-2 positive (G3) (n = 5)** | 93 (231) | 182 (3980) | 1895 (4492) | 1158.5 (2673.5) |
| **Ad5-nCoV/BNT/SARS-COV-2 positive (G4) (n = 2)** | 3.8 (-) | 61.05 (-) | 35.56 (-) | 2090 (-) |
| **p- value\*** | 0.030 | 0.313 | 0.028 | 0.050 |

Data are presented as median (IQR). Kruskal Wallis test was performed for comparisons.
Comparison between the initial and final measure; A p-value $\leq 0.05$ was considered statistically significant.

## Discussion

This study compared the quantitative SARS-CoV-2 spike 1-2 IgG antibody titers after the immunization with the Ad5-nCoV and after combining Ad5-nCoV and BNT. The group immunized with a heterologous vaccine scheme reported higher antibodies titers with no serious adverse events when vaccines were applied ninety days apart. In addition, patients with previous SARS-COV-2 history reported more elevated antibodies titers as well.

Previous studies compared the SARS-CoV-2 spike 1-2 IgG antibodies titers between a single dose of an adenovirus-vector (Oxford/AstraZeneca), or mRNA (BNT or mRNA-1273) vaccine to analyze the efficacy of a one-shot vaccine. In their study, Alexandra J. Spencer et al. reported that a single dose of vaccine (either adenovirus-vector or mRNA) had lower antibody titers than two doses of mRNA vaccine or a heterogeneous scheme with adenovirus-vector and mRNA [3]. Some authors agreed that a heterologous vaccine regimen leads to a strong induction of antibodies. Such is the case of Tina Schmidt et al., who reported that IgG titers of an adenovirus-vectored vaccine and mRNA similar heterologous regimens were a significant improvement on the antibodies’ titers, being approximately tenfold higher than those obtained after homologous vector vaccination [13]. Our study is consistent with the increase of IgG levels in the group where we used a heterologous regimen. The groups that received a BTN boost reported a much higher antibody titer than a single dose of an adenovirus-vector vaccine.

Studies about heterologous vaccination were started because of the concern of the adverse effects of the vaccines. For example, in March 2021, in Germany, the Oxford/AstraZeneca vaccine administration was suspended due to the concern of vaccine-induced cerebral venous thrombosis and thrombocytopenia syndrome, primarily in younger women [13]. This resulted in implementing a strategy of a second dose with an mRNA vaccine in people who received an adenoviral-vector-vaccine as the first dose [14]. This recommendation was for people younger than 60 or 65 years in several European countries [15]. Annabel A Powell et al. refers to the clinical advice because severe reactogenicity after the first dose is the most common reason for receiving a heterologous vaccine regimen [14]. However, in our study, the motivation to start a new vaccine schedule was due to the patients’ doubt about vaccine effectiveness. They did not receive any previous medical advice; the decision was taken on their own.

Nonetheless, the adenovector-based vaccines, such as Ad5-nCoV and Oxford/AstraZeneca, are not the only ones to struggle with the adverse reaction rates. According to Qian He et al., mRNA vaccines, such as BTN, had raised concerns due to the high incidence of adverse reactions; that is why the heterologous regimen had been promoted. A study comparing a single-dose SARS CoV-2 vaccine, priming with adenovector vaccines followed by inactivated vaccines, recombinant protein, or mRNA vaccines, concluded that sequential immunization with a heterogeneous regimen could potentially mitigate the adverse effects of mRNA vaccines [16]. Tina Schmidt et al. compared the reactogenicity of a heterologous regimen with an mRNA boost. The results showed that both local and systematic events were less severe and well-tolerated than the homologous mRNA regimens [13].

We evaluated the vaccine’s adverse events and their perception after receiving each immunization. There were more adverse events reported after the Ad5-nCoV than the BNT. The most-reported reaction after the Ad5-nCoV was headache, tiredness, and pain at the injection site, and their severity was classified mainly as very low, similar to previously reported [17]. On the other hand, the number of adverse events reported after the BNT boost shot was less, being the most frequent: pain at the injection site, tiredness, and headache, and their severity perception was low. Our study concluded that the number of adverse effects with BNT boost is less, but the perception of its severity was milder with Ad5-nCoV. Other studies had shown similar data, where heterologous schedules with a BNT boost had reported more intense reactogenicity than those who received the homologous-counterparts [18,19].

To the best of our knowledge, there are no previous studies where one dose of Ad5-nCoV and a BNT boost have been studied. We think this study is valuable because it shows a good response in quantitative SARS-CoV-2 spike 1-2 IgG antibody titers against SARS-COV-2 with no severity in adverse reactions. It also shows the difference in the antibodies ‘ levels between patients with a history of SARS-COV-2, demonstrating a higher level as previously published with other types of vaccines [20].

The approval of heterologous vaccination could be an opportunity to create more flexible vaccination programs, which is a significant issue in countries with scarce vaccine access or countries where different vaccines are available at other times, besides the appearance of new SARS-CoV-2 variants [19]. In addition, two-dose regimens could also be challenging to complete if they experienced anaphylaxis or any severe reaction after the first dose, leading to recommending a different vaccine for their second dose [14].

A limitation of this investigation is the sample size. So further prospective studies must recruit a more significant sample. Another limitation was the time between both vaccines. It could have more relevance to justify the time lapses, but it was established according to the patients’ access to another vaccine.

## Conclusion

Patients who received a one-shot vaccine reported lower antibody titers than those with a heterologous regimen, with less severe adverse reactions in the short term. Nevertheless, it is crucial to keep following their evolution for a long time to acknowledge its security and efficacy.

The most supplied vaccines worldwide are BNT and Oxford/AstraZeneca, which explains most investigations spin around them. This study analyzed a heterologous regimen based on BNT and Ad5-nCoV, a less frequent vaccine. It may be considered an original investigation because no similar studies were found when this article was written. The importance of pursuing deeper investigations about every SARS-CoV-2 vaccine is mainly based on the fluctuations in logistical suppliance. The antibodies measurement had been considered a favourable rate that mirrors the efficacy of the vaccines; it will be interesting to analyze whether it relates to similar efficacy.

## Data Availability

data available upon request

---

**Figure 1.**
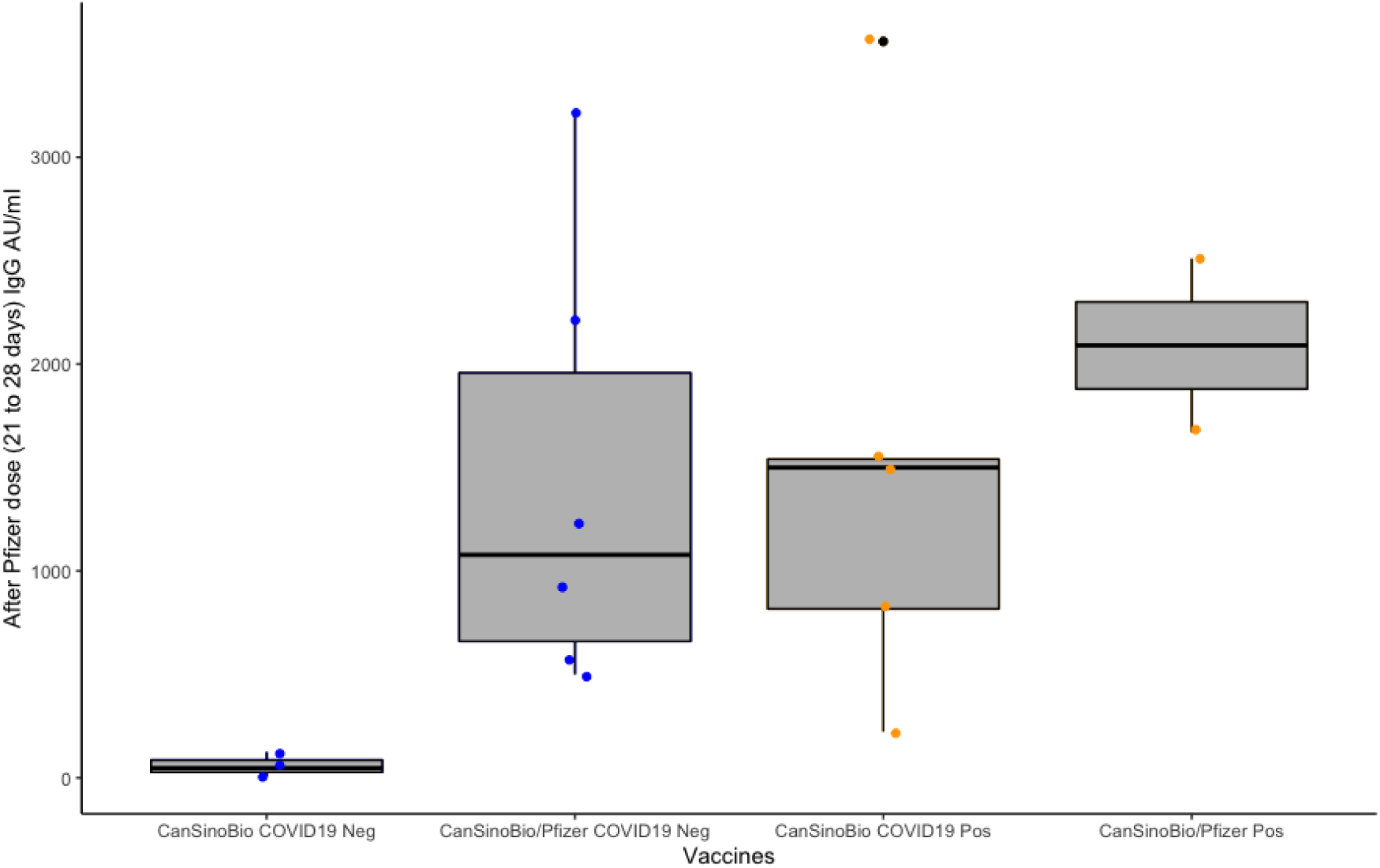
Anti-SARS-CoV-2 spike 1-2 IgG antibodies titers compared between groups. Boxplot that represents the IgG antibodies titers between the four different groups: patients vaccinated with Ad5-nCoV with no known history of SARS-COV-2, patients vaccinated with Ad5-nCoV and the first shot of BNT with no known history of SARS-COV-2, patients vaccinated with Ad5-nCoV with a known history of SARS-COV-2, and patients vaccinated with Ad5-nCoV and the first shot of BNT with a known history of SARS-COV-2 after 21-28 days of BNT boost.

